# Trajectories of mHealth-tracked mental health symptoms and their predictors in chronic pelvic pain

**DOI:** 10.1101/2024.09.25.24314368

**Authors:** Emily L. Leventhal, Nivedita Nukavarapu, Noemie Elhadad, Suzanne Bakken, Michal Elovitz, Robert Hirten, Jovita Rodrigues, Matteo Danieletto, Kyle Landell, Ipek Ensari

## Abstract

**Background.:** Female chronic pelvic pain disorders (CPPDs) affect 1 in 7 women worldwide and are characterized by psychosocial comorbidities, including reduced quality of life and 2-10 fold increased risk of depression and anxiety. Despite its prevalence and morbidity, CPPDs are often inadequately managed with few patients experiencing relief from any medical intervention. Characterizing mental health symptom trajectories and lifestyle predictors of mental health is a starting point to enhancing patient self-efficacy in managing symptoms. Here, we investigate the association between mental health, pain, and physical activity (PA) in females with CPPD and demonstrate a method for handling multi-modal mobile health (mHealth) data.

**Method.:** The study sample included 4,270 person-level days and 799 person-level weeks of data from CPPD participants (N=76). Participants recorded PROMIS global mental health (GMH) and physical functioning, and pain weekly for 14 weeks using a research mHealth app, and moderate-to-vigorous PA (MVPA) was passively collected via activity trackers.

**Data analysis.:** We used penalized functional regression (PFR) to regress weekly GMH-T (GMH-T) on MVPA and weekly pain outcomes, while adjusting for baseline measures, time in study, and the random intercept of the individual. We converted 7-day MVPA data into a single smooth using spline basis functions to model the potential non-linear relationship.

**Results:** MVPA was a significant, curvilinear predictor of GMH-T (p<0.001), independent of pain measures and prior psychiatric diagnosis. Physical functioning was positively associated with GMH-T, while pain was negatively associated with GMH-T (β=2.24, β=-1.16, respectively; p<0.05).

**Conclusion:** These findings suggest that engaging in MVPA is beneficial to the mental health of females with CPPD. Additionally, this study demonstrates the potential of ambulatory mHealth-based data combined with functional models for delineating inter-individual and temporal variability.

## Introduction

Described as a “neglected reproductive health morbidity,” chronic pelvic pain (CPP) is a highly debilitating condition that affects between 5.7% and 26.6% of women worldwide.^1–3^ CPP, which encompasses complex CPP disorders (CPPDs) such as endometriosis, adenomyosis, and fibroids, is characterized by non-cyclic pain in the pelvis or abdomen that lasts for at least 6 months and leads to functional disability or the necessity for medical intervention.^3–5^ Its severity is underscored by its associated physical, psychological, and emotional, and social consequences.^4,5^

The strong psychosocial impact of CPPDs contributes to their morbidity. For example, individuals with CPPDs are more likely to experience reduced quality of life, emotional well-being, productivity, and sexual function compared to the general population.^4^ Additionally, CPPD patients have a significantly higher risk of comorbid psychiatric disorders.^4,6,7^. For example, individuals with CPPDs have been reported to experience depressive disorders at a prevalence of 2 to 10 times that of the general population and anxiety disorders 3 to 6 times that of the general population.^4^ Because chronic pain is tightly linked to mental health problems, investigation of potentially modifiable predictors of mental well-being in individuals with CPPDs may be a starting point for comprehensively managing and treating CPPD patients.^8^

Despite its prevalence and burden, CPP is often inadequately managed, with less than half of patients experiencing pain relief from any medical treatment.^6,9^ Patient self-management, which encompasses active efforts to manage pain and its effects on physical and emotional function, is a common chronic pain care model intervention, and it has been associated with significant improvement in symptoms.^10,11^ Further, Center for Disease Control (CDC) guidelines state that non-opioid and non-pharmacologic therapies should be prioritized for chronic pain management.^12,13^ Non-pharmacological self-management strategies, especially those that target mental health outcomes of CPPD patients, are needed for effective personalized treatment of CPPD.

Physical activity (PA), and exercise, defined as planned, structured, and repetitive PA with the goal of improved health or fitness, have been demonstrated to be effective pain self-management for both reducing pain severity and improving psychological function in chronic pain patients.^11,14^ Experts recommend that chronic pain patients exercise on a regular schedule on the premise that avoiding activity during pain and increasing intensity later may lead to pain flares.^4^ Importantly, exercise is a modifiable behavior that can also improve pain self-efficacy, defined as the confidence in one’s ability to function effectively while in pain, which is associated with improved quality of life.^15,16^ Further, for chronic pain patients with comorbid psychiatric conditions, exercise may improve mood, depression, and anxiety symptoms.^4^ A previous study with individuals with endometriosis estimated a small but statistically significant favorable effect of exercise on pain severity.^17^ However, this study relied on self-reported exercise, which is limited in its ability to capture more granular PA parameters (eg, step counts, intensity-level).^17^ While most of the evidence connecting PA to psychosocial improvement has been from other chronic pain conditions, yoga has previously been demonstrated to be efficacious for improving pain and quality of life for patients with endometriosis.^4^ The impact of broader PA on mental health in patients with CPP specifically remains to be investigated, with a focus on using longitudinal data to capture potentially meaningful trends over time.

CPPDs and their symptomatic patterns are notably heterogeneous in clinical presentation both between patients and within-individuals over time.^18^ Capturing these fluctuations under ecologically valid circumstances can help improve our understanding of the dynamic unfolding of these symptoms and their potential predictors. In the context of health behaviors such as PA, data from mobile health (mHealth) technologies (eg, smartphone apps, trackers) combined with longitudinal analytic techniques can help elucidate symptom associations with psychosocial outcomes in CPP.^7,18^ For example, there may be non-linear associations and cumulative effects in these longitudinal data that are not possible to capture via linear modeling approaches. In sum, flexible techniques can be particularly useful when considering variables that differ in sampling frequencies, missingness patterns, modality, and temporal complexity, which is often the case with mHealth data.

Functional regression models, which are a part of the family of generalized additive models (GAMs), constitute one such approach.^19^ In a functional regression framework, the entire data curve is considered as the unit of analysis, instead of discrete data points in a set of longitudinal data. This is particularly useful for handling PA data from wearables, rather than aggregating multiple data points per individual,^20^ as they allow investigating the associations between scalar and functional variables with different time intervals. One example of a scenario relevant to this study is consideration of continuous or daily PA data with weekly self-reported survey data, in a repeated-measures design. This results in a data structure where each weekly questionnaire corresponds to 7 days of PA data leading up to the survey data. A functional regression model considers the PA data as a weekly data curve rather than aggregating the entire week into a summary score and thus preserves the temporal pattern within the data. This can reveal important information that may be lost otherwise, such as periods of inactivity or bursts of activity, which could be related to mental health.^4,21^

Accordingly, this study aims to characterize the patterns of association between self-reported mental health symptoms and their predictors in CPPDs, with a focus on modifiable lifestyle factors. Specifically, this overall aim includes investigation of 1) between- and within-individual fluctuations in weekly self-reported mental health, and 2) possible modifiable and trait predictors of weekly mental health. We hypothesized that there would be significant variability in the mental health both between and within individuals and that PA would be a positive non-linear predictor of mental health.

## Methods

### Study Design and Procedures

The study design and procedures were approved by the IRB of the Icahn School of Medicine at Mount Sinai (ISMMS; IRB# STUDY-22-01002). This is a secondary analysis of the data from an ongoing larger study that aims to design, develop, and evaluate CPPD-specific mHealth measures from patient generated health data with high complexity and temporality using non-linear distributed lag and functional data modeling (NIH/NICHD: R01HD108263). It uses an observational study design to collect 90 days of data on patient self-tracked symptoms via a research mHealth app (ehive^22^) and passively collected activity data using activity trackers from participants. All participants used the ehive research study app for providing the baseline and weekly data on overall health, symptoms, well-being and health behaviors, as well as for receiving prompts and reminders about the study.^22^ Participants were instructed to wear a Fitbit for the duration of the study.

### Study Sample

The study sample included individuals who met the following eligibility criteria for the parent study: 1) females who menstruate currently, between the ages of 18 and 64, 2) self-reported CPPD based on clinician diagnosis, 3) experience of CPP for at least 6 months, and 4) ability to read and write in English. Exclusion criteria include: 1) current pregnancy, a birth in the past 6 months, or planning pregnancy during the months of the study and 2) major diseases or comorbidities (eg, active cancer, acute coronary syndrome within the past 3 months) that might confound the outcomes of the primary pelvic pain-related condition. Participants were recruited from all campuses of the Mount Sinai Health System (MSHS) and Columbia University Irving Medical Center (CUIMC) via email advertisements and on the myChart by EPIC mobile app for MSHS patients.

### Enrollment

Interested patients reached out to the study coordinator at Mount Sinai for screening and enrollment, after which they were onboarded and oriented to the study app and data collection protocols. All participants were mailed a Fitbit Inspire 2 device and instructed to use for the duration of the study (90 days). Participants were remunerated $15 for every 2 weeks of data collection and $20 for the final week (ie, up to $120 in total for completing 90 days of data collection). All participants provided informed consent prior to enrolling in the study.

### Study Measures

#### Primary Outcomes

Self-reported mental health was assessed every week using the PROMIS Global Mental Health Questionnaire (GMH; 2a, v1.2).^23^ The GMH includes 2 questions: 1) “In general, how would you rate your mental health, including your mood and your ability to think?” 2) “In general, how would you rate your satisfaction with your social activities and relationships?” Both questions have a 5-point multiple choice response scale (1-not at all, 5-very much) and the responses are added to compute the total score on the GMH (range 2-10). Higher scores represent better mental health.^23^ The two-item GMH survey provides a brief measure of mental health that has been found to be both reliable and have construct validity.^23^ Scores from the GMH survey have been positively associated with other self-reported outcomes including overall quality of life and physical functioning, and negatively correlated with fatigue, anxiety, anger, depressive symptoms, and chronic conditions (e.g,. liver disease, kidney disease, hypertension, etc.).^23^ We converted raw GMH scores to population-standardized GMH scores (T-scores) according to the PROMIS Global Health scoring manual by standardizing the raw total score to a mean of 50 and a standard deviation (SD) of 10.^24^ GMH T-scores (GMH-T) are further categorized as excellent (>55), very good (48-55), good (40-47), fair (29-39), and poor (<29).^25^

#### Predictors

##### Physical activity

Daily minutes of moderate-to-vigorous intensity PA (MVPA) and step counts were obtained from the wrist-worn Fitbit devices. Participants were instructed to wear their devices continuously for the study duration. The study app (ehive) allows the user to link their account with their Fitbit device,^22^ which enables regular daily data synching on the backend of the app. Fitbit uses its proprietary algorithms for detection of step counts and activity intensities. We collected 6,341 days of physical activity data for 78 participants. For wear time validation, we relied on the commonly used standard “10-hour minimum wear” rule, in which a valid day is defined as at least 10 hours of non-zero activity counts.^26–28^ Ten hours of wear has been shown to be sufficient to estimate total daily physical activity during non-sleep time.^29^ There were 4,301 valid days of Fitbit data for 76 participants. Days with unrealistically low activity counts (eg, <500 steps in a day; n=14) were removed in accordance with similar cutoffs that have been used in the past to define a valid day, although we used a more conservative cutoff.^26,27^ This resulted in 4,287 days of physical activity data for 76 participants. If there were more than 7 days of Fitbit data in between survey responses (ie, if a participant waited more than 7 days before completing the next survey), we only considered the first 7 days of Fitbit activity data to avoid sparsity in the penalized functional regression (PFR) model (described below). 77 days of activity data measured more than 7 days after a survey response were removed for this reason. The final dataset had 4,270 days of data for 76 participants.

##### Physical functioning

Weekly physical functioning scores were measured using the PROMIS physical function survey (4a, v1.0).^30^ Physical functioning is the self-reported capability of performing everyday physical activities. The score evaluates functioning of upper extremities, lower extremities, central regions, and activities of daily living. The 4-item PROMIS survey assesses the extent to which individuals find difficulty with physical tasks (5-without any difficulty to 1-unable to do). Scores range from 4 to 20, with higher scores indicating better physical functioning. We used the physical functioning T-scores in the analyses, which are standardized to a mean of 50 and a SD of 10 based on a representative population distribution.^30^

##### Pain

We measured weekly pain levels using the VAS pain intensity item from the short-form McGill Pain Questionnaire (MPQ-VAS).^31^ The MPQ-VAS asks participants to rate the intensity of their present pain intensity on a scale of 0 (no pain) to 100 (worst imaginable pain).^32^ This type of VAS-based pain assessment is commonly used as a standard practice in clinical settings to evaluate patient pain status and treatment outcomes.^33,34^

##### Other covariates

Data on personal demographics and general health were collected via a baseline questionnaire on the ehive app. We collected age, marital status, ethnicity, and employment status from the demographics survey. In addition, we used prior psychiatric diagnosis (“Have you ever been diagnosed with a psychiatric diagnosis by a provider?”) as a covariate from the general health survey.

## Data analysis

### Descriptive and bivariate analyses

First, we performed descriptive analyses and investigated bivariate associations between the weekly-measured survey items. Given the repeated-measures design, we use both person-level means (ie, a participant’s mean score across the 14 weeks) and overall sample means (ie, mean of means) where necessary to report the overall study average scores from the daily (ie, steps, MVPA) and weekly (ie, pain, physical functioning T-score, GMH-T) measures. To analyze the GMH-T, we converted the mean GMH-T for each participant to its corresponding GMH category (eg, fair, good, excellent, etc.), and computed the percent of participants in each category.^25^ To evaluate sample GMH-T and physical functioning T-scores against known population means, we used a one-sample T-test to compare the sample means to the population means. We then computed repeated-measures correlations between GMH-T, physical functioning, MPQ-VAS, and the sum of MVPA over 7 days using the rmcorr R package, which evaluates the within-individual association of paired measurements taken two or more times longitudinally.^35^

### Multivariable regression analysis of GMH predictors

To investigate the potential predictors of GMH-T scores at the week level, we implemented PFR modeling using the R refund library.^20^ PFR models are flexible in numerous ways that are particularly useful for the data in this study. First, they allow for entire data curves to be units of analysis as opposed to individual data points. Next, they accommodate different sampling intervals in the outcomes vs predictors, ie, week-level outcome (eg, GMH-T) and week-level (eg, pain, physical functioning) and day-level (eg, MVPA) predictors. Instead of aggregating multiple day-level MVPA values for each week, this feature of the PFR allows for the preservation of temporal variability in MVPA over a week. Third, it allows specification of random intercepts (ie, individual participants), which is useful for both accommodating a repeated measures design and for investigation of potential between-vs within-individual variability in the outcome of interest (ie, GMH-T scores).

We regressed GMH-T on MVPA while considering MPQ-VAS, PROMIS physical functioning, age, marital status, employment status, and prior psychiatric diagnosis. We further adjusted for time in study using month-level cyclical encoding, in which each date is mapped into a cyclic coordinate system using sine-cosine waves and allows the models to infer the distance between dates based on their sine-cosine coordinates. We converted 7-day MVPA data into smooths with up to 7 knots using the tensor product basis function^36^ to model the potential non-linear relationship between GMH-T and daily PA. We similarly included the time covariate as a functional smooth with up to 7 knots.^20^ We scaled MPQ-VAS, PROMIS physical functioning, and age by mean-centering each variable and dividing by its standard deviation. We included participant and week in study as random effects. Finally, other categorical variables (ie, psychiatric diagnosis, employment status, and marital status) were included as person-level linear covariates.^20^ We used a generalized additive model as the fitter to estimate the model and restricted maximum likelihood as the smoothing parameter estimation method, which are the default recommended methods for the function.^20^

## Results

### Study sample

Participants (n=76) provided 799 weeks of survey and 4,270 days of activity data in total for analysis. Participants had a mean age of 35 years and were mostly employed (76%). Most participants identified as White (42%) or Hispanic or Latino (17%). In our sample, 28% had at least one prior diagnosis of a psychiatric condition, including anxiety and mood disorders (Table 1). The CPPD diagnoses included endometriosis (N=51), adenomyosis (N=1), uterine fibroids (N=2), interstitial cystitis (N=1), inflammatory bowel syndrome (3), and inflammatory pelvic dysfunction (N=1).

**Table 1.**
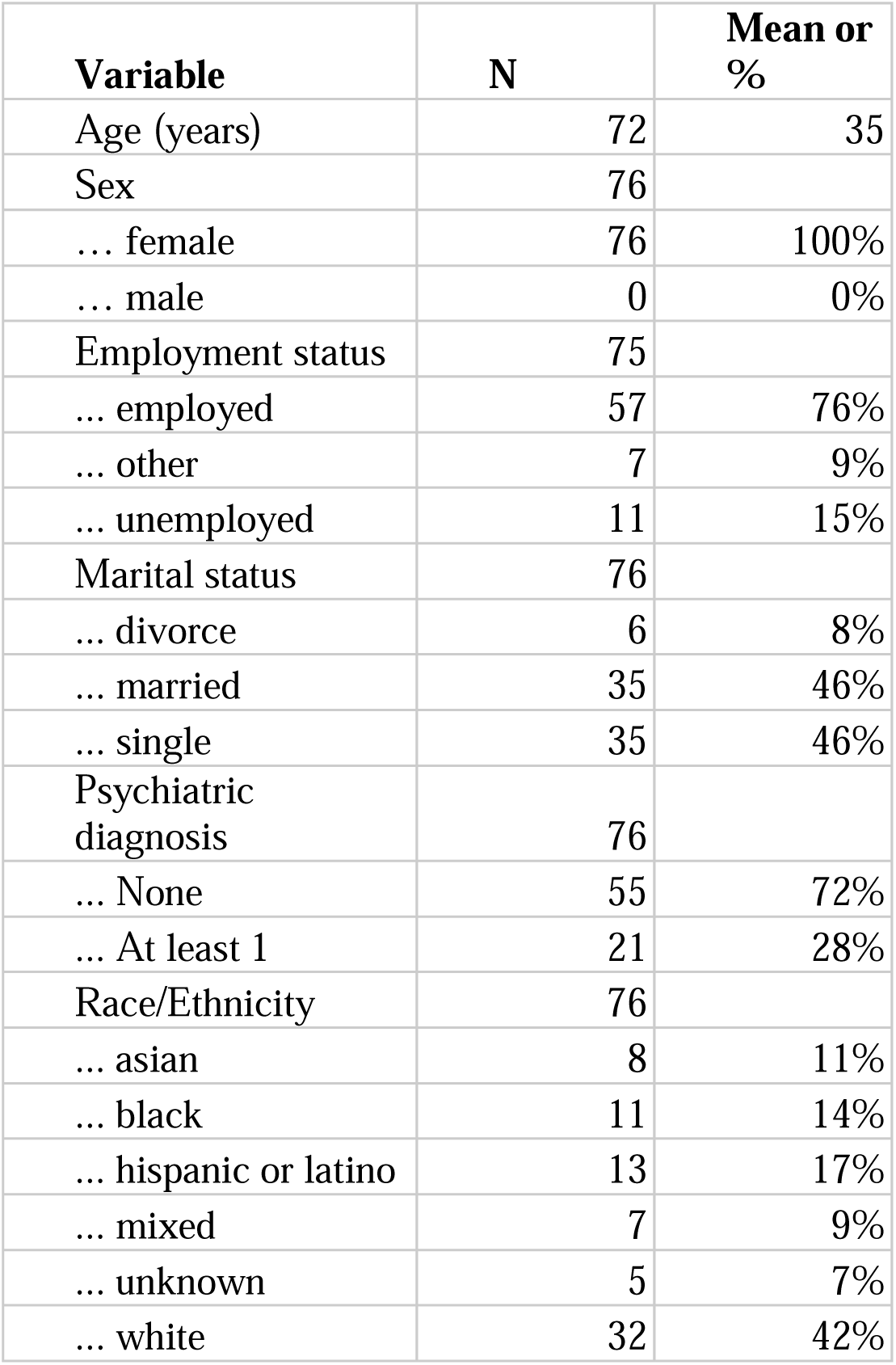
Study sample demographics.

| **Variable** | **N** | **Mean or %** |
| --- | --- | --- |
| Age (years) | 72 | 35 |
| Sex | 76 |  |
| ... female | 76 | 100% |
| ... male | 0 | 0% |
| Employment status | 75 |  |
| ... employed | 57 | 76% |
| ... other | 7 | 9% |
| ... unemployed | 11 | 15% |
| Marital status | 76 |  |
| ... divorce | 6 | 8% |
| ... married | 35 | 46% |
| ... single | 35 | 46% |
| Psychiatric diagnosis | 76 |  |
| ... None | 55 | 72% |
| ... At least 1 | 21 | 28% |
| Race/Ethnicity | 76 |  |
| ... asian | 8 | 11% |
| ... black | 11 | 14% |
| ... hispanic or latino | 13 | 17% |
| ... mixed | 7 | 9% |
| ... unknown | 5 | 7% |
| ... white | 32 | 42% |

### Descriptive and bivariate analyses

The overall sample means of the scores from the daily and weekly measures are reported in Table 2. Thirty-nine percent of the participants, on average, reported scores that corresponded to “fair” mental health, with another 39% of the participants on average reporting “good” mental health (Table 2). The mean GMH-T was 42.166 (95% CI: 40.363-43.969), which is 7.83 SDs below the population mean (ie, M=50, “very good”)^23^ and significantly different (*t*=-8.658, *p* < .001; Figure 1). The mean physical functioning T-score was 45.19 (95% CI: 43.52-46.853), which is 0.48 SDs below the population mean (ie, M=50; Figure 1; *t* = -5.758, *p* < .001).

**Table 2.** Average weekly and daily measures across the study. The average was taken of the participant means for each repeated measure.

| **Variable** | **N** | **Mean or %** |
| --- | --- | --- |
| Mean MVPA | 76 | 38 |
| Mean steps | 76 | 8313 |
| Mean MPQ-VAS | 75 | 34 |
| Mean phys. func. T-score | 73 | 45 |
| Mean GMH T-score | 75 | 42 |
| Mean GMH T Category | 75 |  |
| ... Poor | 3 | 4% |
| ... Fair | 29 | 39% |
| ... Good | 29 | 39% |
| ... Very Good | 8 | 11% |
| ... Excellent | 6 | 8% |

**Figure 1.**
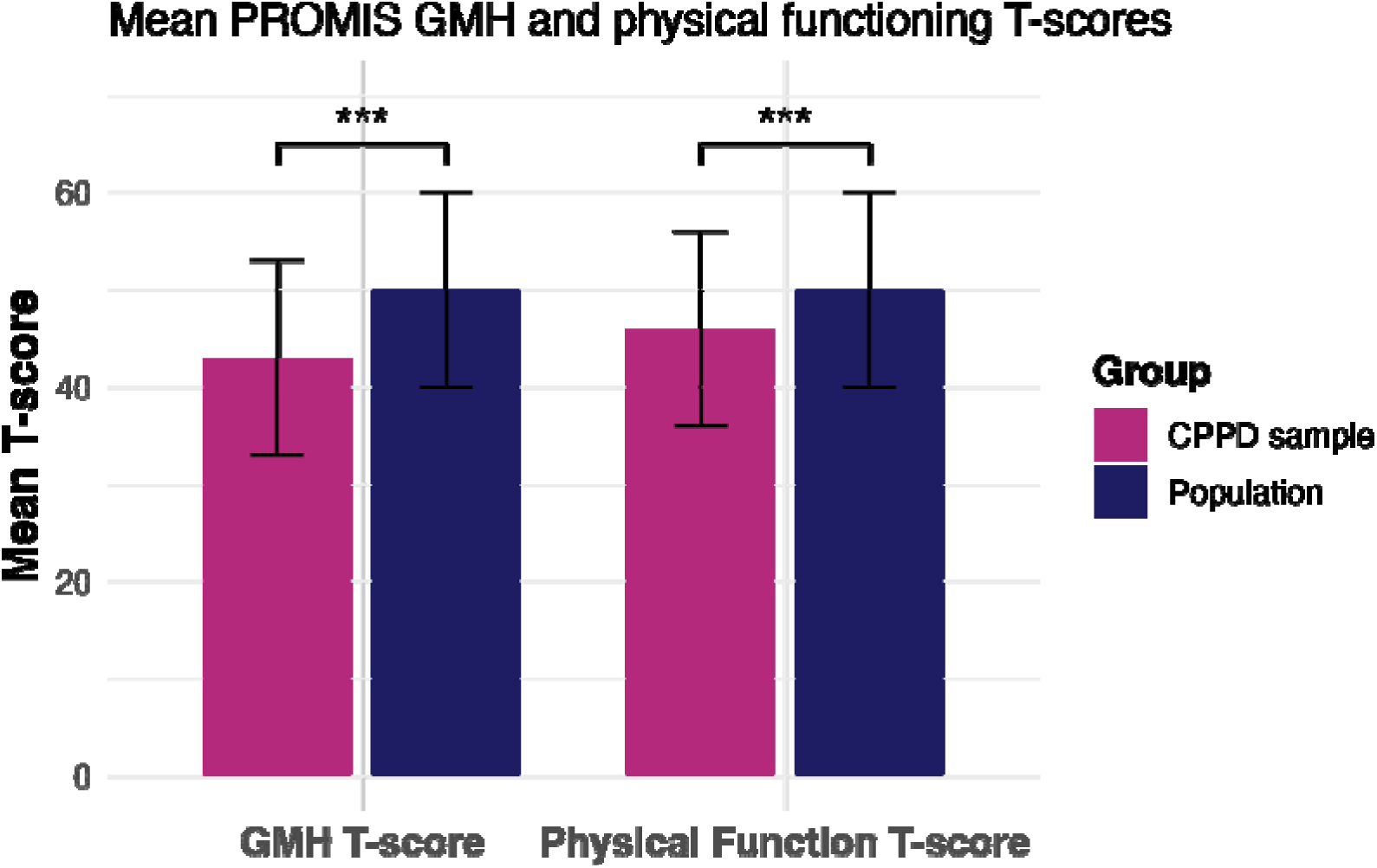
- - - - - =42.166, 95% CI: 40.363-43.969, M=50, *t*=-8.658, *p* < .001) and physical - =45.19, 95% CI: 43.52-46.853, M=50, *t* = -5.758, *p* < .001) means were significantly different than the general population.

To characterize the PA patterns in the sample, we compared participants’ activity levels to the published recommendations and CDC/HHS PA guidelines for adults with respect to steps and MVPA.^37,37–39^ On average, participants accumulated 8,313 steps and 38 minutes of MVPA per day (Table 2). Forty-three percent of the sample engaged in fewer than 7,500 daily steps, which is the lower threshold recommended for being considered “sufficiently active” (Figure 2a). ^38,39^ Similarly, 40.9% accumulated fewer than 150 minutes of weekly MVPA recommended by the PA Guidelines (Figure 2b).^40^

**Figure 2.**
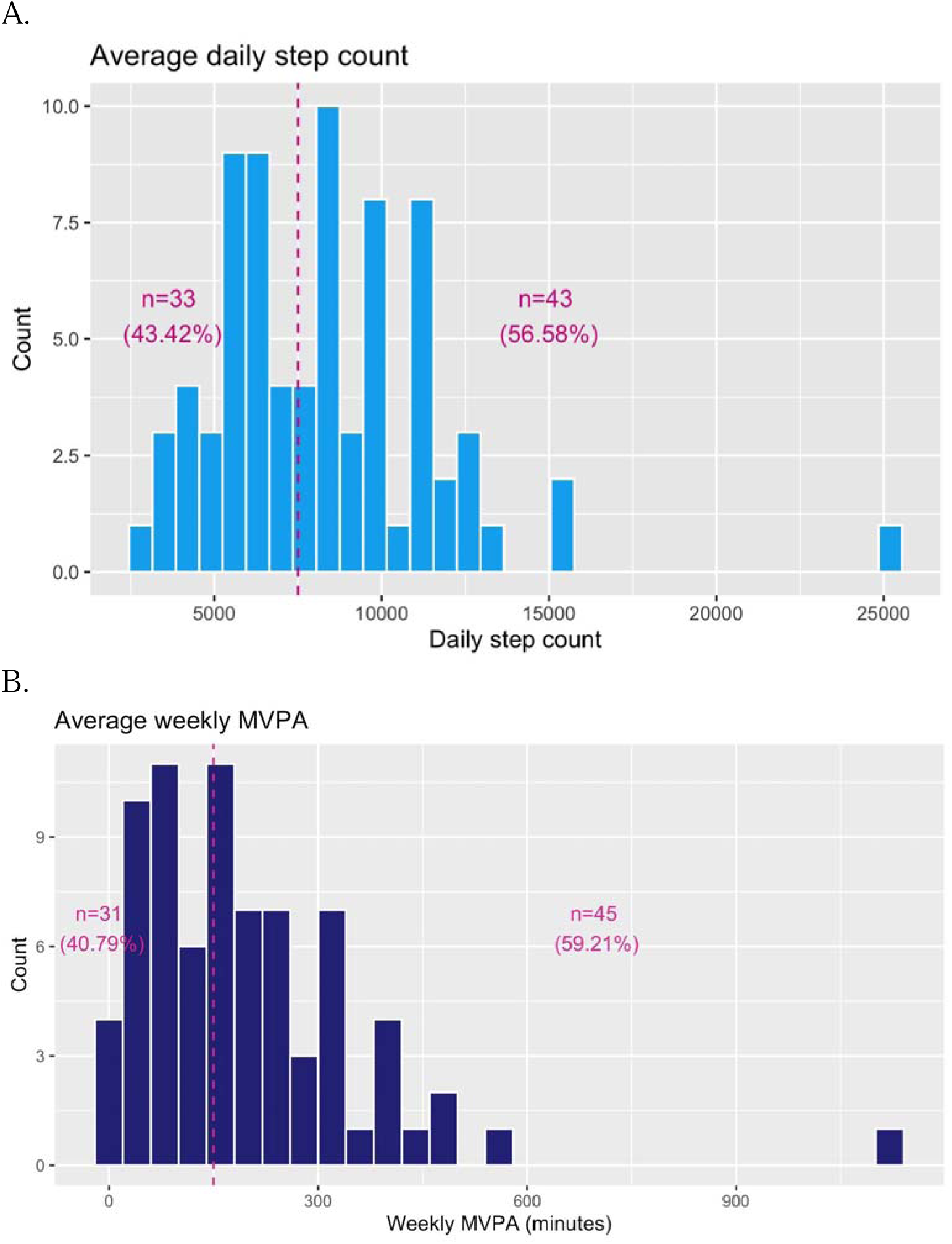
Mean participant **A)** daily step count and **B)** mean weekly MVPA minutes compared to nationally recommended activity levels. The y-axis represents the number of participants. Dashed lines represent the recommended levels (7500 steps, 150 MVPA minutes). The values represent the number of individuals who fell above and below these nationally recommended values.

To inspect the bivariate associations between weekly measures, we computed repeated measures correlations between GMH-T and the other variables. GMH-T were positively correlated with weekly MVPA (*p*<.05), and physical function T-score (*p*<.01), while they were negatively correlated with MPQ-VAS (*p*<.001; Figure 3). Weekly MVPA was additionally positively correlated with physical functioning T-score (*p*<.05) but was not significantly correlated with MPQ-VAS.

**Figure 3.**
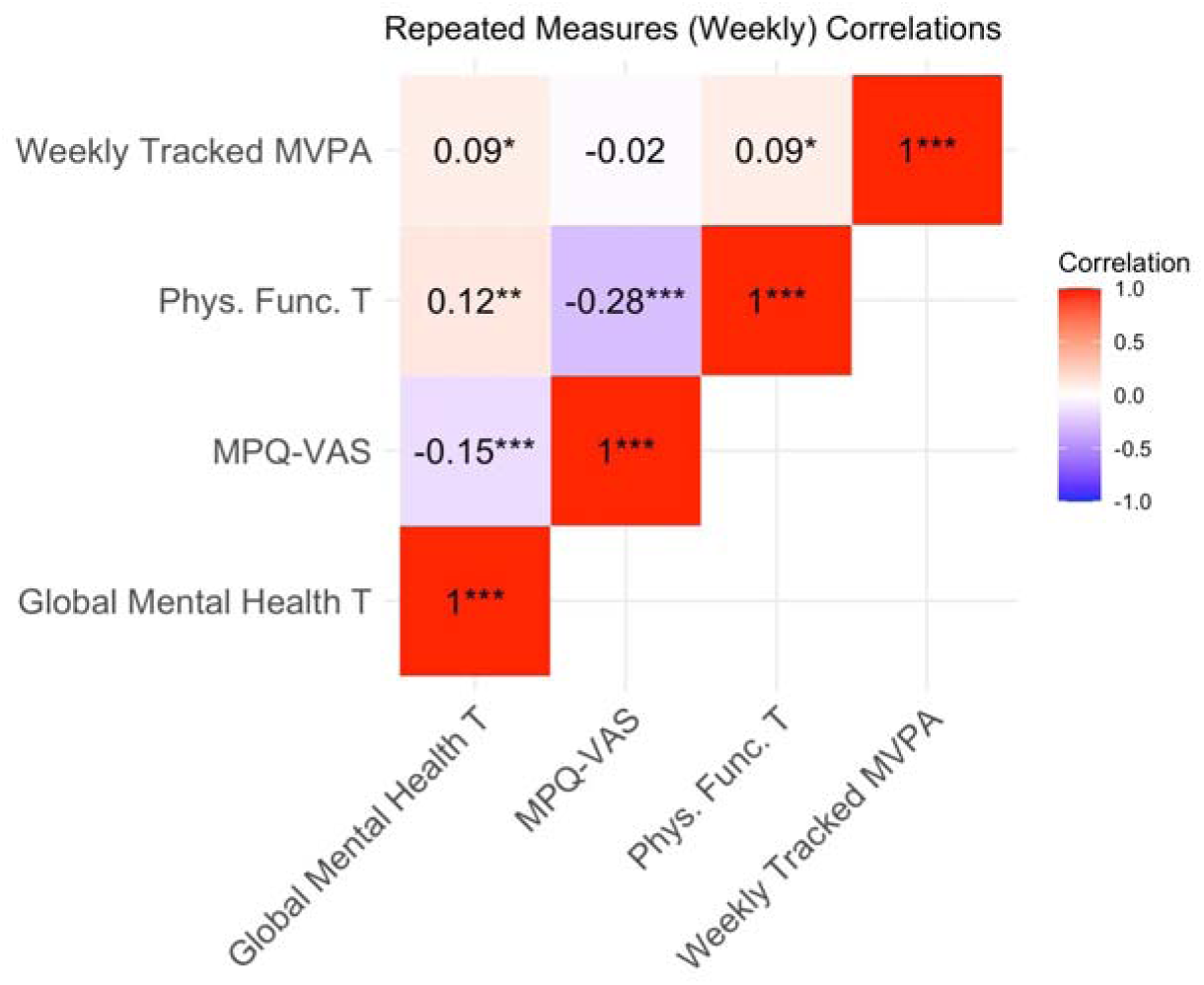
Repeated measures correlations for weekly measures. MVPA=moderate-to-vigorous physical activity; Phys. Func. T = physical functioning T-score; MPQ-VAS=McGill Pain Questionnaire-VAS; Global Mental Health T=GMH-T

### PFR model

We fitted a PFR model to the data to investigate cumulative and non-linear effects of MVPA on the weekly GMH-T. The best fitting final model explained 72.6% of the variance in GMH-T (R^2^=0.65). The smooth of MVPA and time on GMH-T indicated a significant non-linear relationship (Table S1; Table S2; edf=2.23, *F*=18.99, *p*<.001). Predicted GMH-T increased with increasing daily MVPA minutes (Figure 4a). Over time, the largest positive effect of MVPA on predicted GMH-T as reported at the end of the week was a few days prior (∼day 4). The positive effect of MVPA on GMH-T reported at the end of the week diminished after day 4, suggesting the positive effects of MVPA lagged by a couple of days. Weekly MPQ-VAS was a significant negative predictor of GMH-T (β=-1.16, SE=0.50, *t*=-2.34, *p*<.05), while physical functioning T-score was a significant positive predictor of GMH-T (Figure 4b; Table S3; β=2.24, SE=0.598, *t*=3.75, *p*<.001). For demographic factors, age was negatively associated with GMH-T (β=-1.20, SE=0.46, *t*=-2.58, *p*<.05), while being employed and married were positively associated with GMH-T (β=4.01, SE=1.09, *t*=3.67, *p*<.001; β=3.60, SE=0.86, *t*=4.20, *p*<.001). Prior psychiatric diagnosis was not a significant predictor of weekly GMH. The random effect of participant was significant (Figure 4c; edf=33.43, *F*=2.76, *p*<0.001). The random effect of week and the cyclically encoded sine and cosine functions of month were not significant.

**Figure 4.**
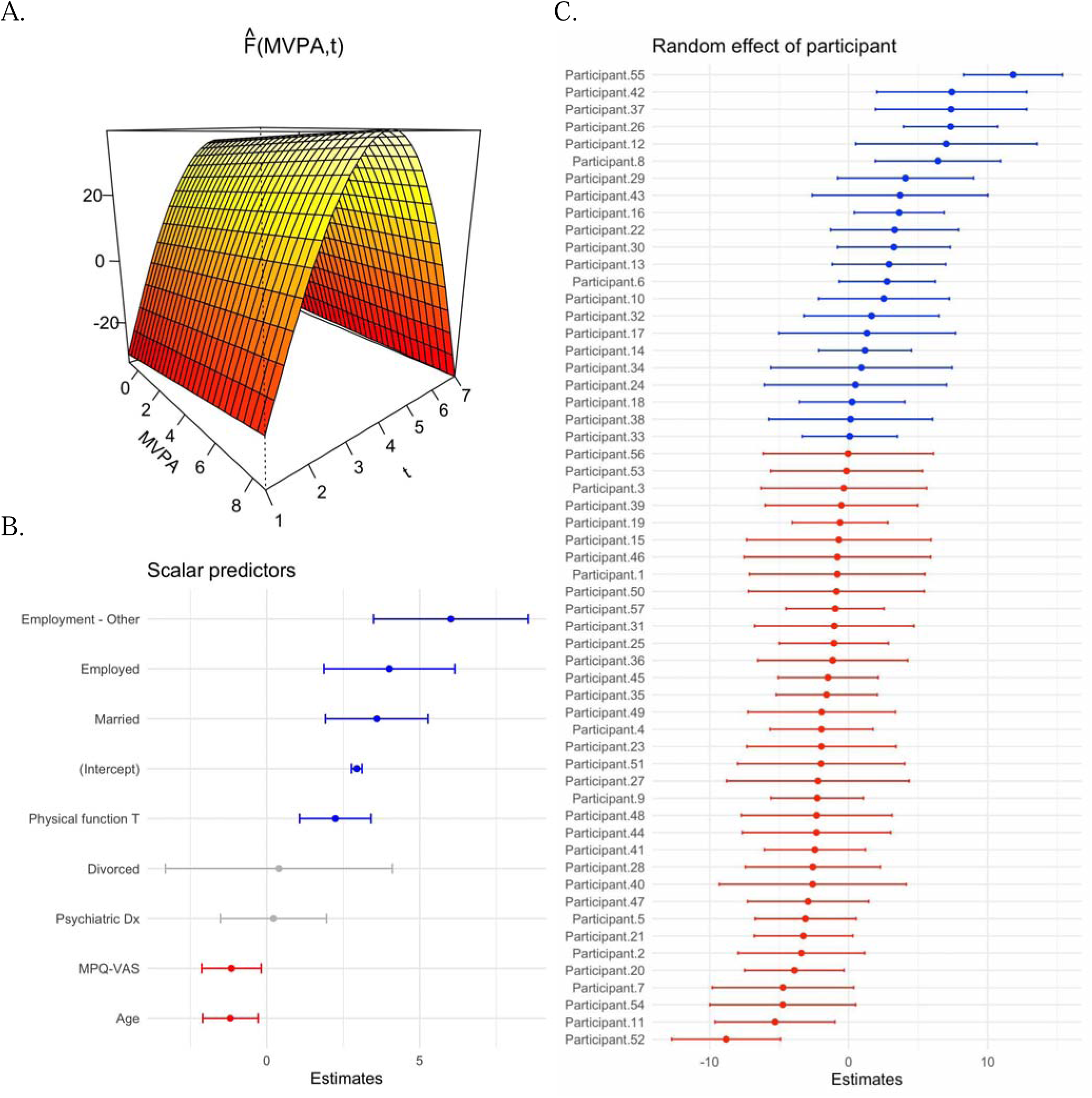
Results from the PFR model. **A)** The smooth effect of MVPA on GMH-T over time. The MVPA axis is scaled. The y-axis represents predicted GMH-T and is scaled according to predicted sample GMH-T mean. **B)** Coefficients and confidence intervals for scalar predictors of the model. **C)** Random effect of participant, with each dot representing predicted mean GMH-T for that participant.

## Discussion

In this study, we leveraged ambulatory mHealth-tracked mental health, pain, and physical activity data to characterize longitudinal self-reported mental health patterns of individuals with CPPDs. Our results indicate a positive, non-linear relationship between PA and mental health, independent of prior psychiatric diagnosis or other pain-related factors, with considerable variability both between and within participants over time. To our knowledge, this study provides the first line of evidence on the positive effect of PA on mental health in females with CPPDs using repeated measures data collected in real time. We further report lower scores of mental health and physical functioning compared to the general population, as well as lower PA levels than those recommended by the PA guidelines.

Our cohort had a 28% incidence of prior psychiatric conditions and lower average global mental health compared to the general population. Chronic pain, and specifically CPPDs, has been established as a strong predisposing factor for psychiatric conditions, due to both the psychosocial impact of chronic pain and common neurobiological vulnerabilities and genetic factors between chronic pain and mood.^4,6,41,42^ CPPD patients with comorbid psychiatric conditions are more likely to incur higher health care costs, experience lower quality of life, endure increased disability, and are more likely to be prescribed opioids.^4^ Additionally, our findings add to the literature documenting the worsened mental health of CPPD patients as a whole compared to the general population.^4,23,42^ In the 2019 National Health Interview Survey, those with chronic pain had a 23.9% prevalence of co-occurring anxiety and/or depression symptoms, whereas the population without chronic pain had a prevalence of 4.9%.^42^ Given the high incidence of psychiatric co-morbidities and the generally low mental health among CPPD patients, it is important to treat mental health as part of comprehensive chronic pain management and continue to determine ways to aid patients to manage their symptoms. As such, here, we investigated how lifestyle factors may modify the association of CPP with poorer mental health outcomes.

Our findings suggest that many females with CPPDs do not reach nationally recommended activity levels, and moreover, that engaging in MVPA is beneficial for the mental health of CPPD patients. The PA levels found in this sample are consistent with previous studies indicating that individuals with CPPDs have lower PA levels,^43^ though data on CPPDs are scarce. One longitudinal study using accelerometers indicated that MVPA negatively mediated the relationship between chronic pain and risk of mental disorders, although this study did not focus on CPP.^44^ Increased MVPA in individuals with chronic pain was associated with decreased anxiety and depression symptoms, whereas light intensity PA did not have this effect.^42^ While previous studies have established the connection between MVPA and mental health in chronic pain, this is the first study to establish the relationship between PA and mental health in the context of CPP by using passively-obtained data from activity trackers.^17^

Our findings further indicate that increased pain is associated with worsened GMH, while increased physical functioning was associated with improved GMH. Though pain and depression or anxiety have been noted to have a bidirectional relationship, there is more evidence that pain is a risk factor for mental health problems than the inverse.^4^ Additionally, a longitudinal study focused on musculoskeletal conditions found that improvements in physical functioning were associated with improved anxiety symptoms, although it was not associated with improved depression symptoms.^45^ The relationship between physical functioning and mental health in CPP has not been well defined to this point, however, one previous longitudinal study on endometriosis reported that functional pain disability did not predict later emotional distress.^46^

With respect to demographic factors as potential predictors, increased age was associated with worsened GMH, while prior psychiatric diagnosis was not a significant predictor. Age may be a proxy for years of experience with the chronic pain condition or severity of the condition. In this study, we did not have a survey item assessing time of initial diagnosis, although this may be possible in the future by linking mobile health studies with electronic health records (EHRs).

Over time, chronic pain may become more difficult to treat due to structural and functional neuroplastic changes that eventually become irreversible and insensitive to treatment.^41^ From a psychosocial standpoint, the economic consequences of health care costs and loss of productivity may accumulate over time.^41^ It will be important to assess how length of time of living with chronic pain impacts mental health in the future. Interestingly, diagnosis with a prior psychiatric condition, including mood and anxiety disorders, was not a significant predictor of GMH. This may suggest that some individuals with prior psychiatric diagnoses may not be actively experiencing symptoms, or alternatively, that this sample has a large number of participants with undiagnosed psychiatric conditions that are actively experiencing symptoms.

We observed substantial between- and within-individual variability in mental health scores in the sample, underscoring the importance of personalized approaches to care. Predicted average GMH-T varied greatly between individuals as shown by the random intercepts. CPPDs are notoriously heterogeneous in pain symptomatology, and it follows that mental health would exhibit similar variability among and within participants.^7^ As such, it is important to use individualized approaches, such as that which may be achieved with mHealth, to comprehensively understand the complexity of CPP. Due to their heterogeneous clinical presentation and differing etiologies, CPPDs are often non-responsive to treatment, and a personalized approach is necessary for the successful management of CPPD. To better understand how to manage the mental health of CPPD patients, we should continue to study modifiable lifestyle factors, as was done here with PA, that may alter the poor mental health outcomes associated with CPP. This study demonstrates the potential of using ambulatory mHealth-based data combined with functional data methods to delineate inter-individual and temporal variability in symptoms of chronic conditions.

There are numerous strengths of this work. First, we focus on a patient population that has been under-studied (ie, CPPDs) and currently still not well-understood as a cluster of disorders with overlapping symptomatology. While endometriosis, the most common underlying primary diagnosis for a CPPD, has been receiving more attention recently, our sample also included those less-studied CPP conditions (eg, adenomyosis, fibroids, inflammatory pelvic disease). Next, implementation of functional data methods and generalized additive modeling using smooths provide robust, flexible approaches for handling the complex patient-generated health data from mHealth technologies. The PFR models in this context facilitate the evaluation of complex relationships between outcomes and their predictors in instances where data sampling frequency differs between the outcomes and predictors, or between different predictors. As mHealth use is becoming more ubiquitous for conducting research, expanding upon the available methods will enable fully harnessing the information from these data. Third, our analyses were based on frequently-sampled prospective data of up to 14 weeks from the study participants. This is a strength of the data design as most studies to date are limited to convenience samples of retrospective data with much less frequency of data points.

Nevertheless, we acknowledge the limitations of this study. Although we had 799 person-level weeks for analysis, 76 participants is a relatively modest sample size in comparison to large, nationally-representative cohort studies. Similarly, the sample was somewhat homogeneous with respect to demographic factors including employment status and education levels. Third, despite our careful inspection of the missing data and implementing cautious filtering criteria to prevent potentially erroneous inference from the data, Fitbit’s proprietary algorithms do not always enable as informed decisions regarding the missing data as do some other devices, such as research grade trackers that allow access to the raw acceleration data. To circumvent these issues, we conducted a series of sensitivity analyses to assess the pattern of missingness in the data, as well as the possible influence of missingness on the model results.

Results (not reported herein) indicated no significant bias, suggesting a missing-at-random (MAR) pattern, or change in model point estimates. Finally, most of the participants had endometriosis as their primary CPPD, therefore we are not able to delineate differences in mental health trajectories among different disorders within CPPD.

## Conclusions

mHealth-enabled direct patient input and passive tracking via wearables enables the capturing of real-world data to improve our understanding of inter-individual and temporal variability in mental health symptoms and factors that may improve mental health. By leveraging patient-tracked mental health and pain outcomes combined with passively-obtained activity data from CPPD patients, we demonstrate a positive, non-linear relationship between PA and mental health in CPP.

## Ethics approval and informed consent

The study was approved by the Institutional Review Board (IRB) of the Icahn School of Medicine at Mount Sinai (IRB# STUDY-22-01002) and all participants provided informed consent.

## Authors’ contributions

All authors contributed significantly to the work presented in this manuscript, including the conception, study design, execution, acquisition of data, analysis and interpretation. Each author reviewed this article and agree to take responsibility for the contents of this article.

## Data Availability

The data collection for the parent grant is currently ongoing. After completion of the active grant period, the data produced in the present study will be made available upon reasonable request to the corresponding author.

## Supplemental Tables

**Table S1.** Smooth predictors of the PFR model.

| Predictor | edf | Ref.df | F | p-value |
| --- | --- | --- | --- | --- |
| t2(MVPA.tmat,MVPA.omat):L.MVPA | 2.23222202 | 2.40665028 | 18.9885386 | 4.0841E-06 |
| s(month_cos.tmat):L.month_cos | 2.00001858 | 2.00003559 | 0.48641865 | 0.61566209 |
| s(month_sin.tmat):L.month_sin | 2.5591852 | 2.8159533 | 0.63612865 | 0.59048914 |
| s(Participant) | 33.4252443 | 57 | 2.75714802 | 1.5399E-06 |
| s(Week) | 5.2859E-05 | 1 | 5.0264E-06 | 0.96005532 |

**Table S2.** Point estimates for smooth terms.

| Predictor | Estimate | SE |
| --- | --- | --- |
| scale(pfr_age) | -1.1946 | 0.4629 |
| t2(MVPA.tmat,MVPA.omat):L.MVPA.1 | 0 | 0.0012 |
| t2(MVPA.tmat,MVPA.omat):L.MVPA.2 | 0 | 0.0012 |
| t2(MVPA.tmat,MVPA.omat):L.MVPA.3 | 0 | 0.0012 |
| t2(MVPA.tmat,MVPA.omat):L.MVPA.4 | 0 | 0.0012 |
| t2(MVPA.tmat,MVPA.omat):L.MVPA.5 | 0 | 0.0012 |
| t2(MVPA.tmat,MVPA.omat):L.MVPA.6 | 0 | 0.0012 |
| t2(MVPA.tmat,MVPA.omat):L.MVPA.7 | 0 | 0.0012 |
| t2(MVPA.tmat,MVPA.omat):L.MVPA.8 | 0 | 0.0012 |
| t2(MVPA.tmat,MVPA.omat):L.MVPA.9 | 0 | 0.0012 |
| t2(MVPA.tmat,MVPA.omat):L.MVPA.10 | 0 | 0.0012 |
| t2(MVPA.tmat,MVPA.omat):L.MVPA.11 | 0 | 0.0012 |
| t2(MVPA.tmat,MVPA.omat):L.MVPA.12 | 0 | 0.0012 |
| t2(MVPA.tmat,MVPA.omat):L.MVPA.13 | 0 | 0.0012 |
| t2(MVPA.tmat,MVPA.omat):L.MVPA.14 | 0 | 0.0012 |
| t2(MVPA.tmat,MVPA.omat):L.MVPA.15 | 0 | 0.0012 |
| t2(MVPA.tmat,MVPA.omat):L.MVPA.16 | 0 | 0.0012 |
| t2(MVPA.tmat,MVPA.omat):L.MVPA.17 | 0 | 0.0012 |
| t2(MVPA.tmat,MVPA.omat):L.MVPA.18 | 0 | 0.0012 |
| t2(MVPA.tmat,MVPA.omat):L.MVPA.19 | 0 | 0.0012 |
| t2(MVPA.tmat,MVPA.omat):L.MVPA.20 | 0 | 0.0012 |
| t2(MVPA.tmat,MVPA.omat):L.MVPA.21 | 0 | 0.0012 |
| t2(MVPA.tmat,MVPA.omat):L.MVPA.22 | 0 | 0.0012 |
| t2(MVPA.tmat,MVPA.omat):L.MVPA.23 | 0 | 0.0012 |
| t2(MVPA.tmat,MVPA.omat):L.MVPA.24 | 0 | 0.0013 |
| t2(MVPA.tmat,MVPA.omat):L.MVPA.25 | 0 | 0.001 |
| t2(MVPA.tmat,MVPA.omat):L.MVPA.26 | 0 | 0.001 |
| t2(MVPA.tmat,MVPA.omat):L.MVPA.27 | 0 | 0.001 |
| t2(MVPA.tmat,MVPA.omat):L.MVPA.28 | 0 | 0.001 |
| t2(MVPA.tmat,MVPA.omat):L.MVPA.29 | 0 | 0.0012 |
| t2(MVPA.tmat,MVPA.omat):L.MVPA.30 | 0 | 0.001 |
| t2(MVPA.tmat,MVPA.omat):L.MVPA.31 | 0 | 0.001 |
| t2(MVPA.tmat,MVPA.omat):L.MVPA.32 | 0 | 0.001 |
| t2(MVPA.tmat,MVPA.omat):L.MVPA.33 | 0 | 0.001 |
| t2(MVPA.tmat,MVPA.omat):L.MVPA.34 | 0 | 0.001 |
| t2(MVPA.tmat,MVPA.omat):L.MVPA.35 | 1.0687 | 0.0317 |
| t2(MVPA.tmat,MVPA.omat):L.MVPA.36 | 0.0002 | 0.1151 |
| t2(MVPA.tmat,MVPA.omat):L.MVPA.37 | -0.123 | 0.0037 |
| t2(MVPA.tmat,MVPA.omat):L.MVPA.38 | 0.0028 | 0.115 |
| t2(MVPA.tmat,MVPA.omat):L.MVPA.39 | -1.3998 | 0.0415 |
| t2(MVPA.tmat,MVPA.omat):L.MVPA.40 | 0.0332 | 0.1141 |
| t2(MVPA.tmat,MVPA.omat):L.MVPA.41 | -0.2206 | 0.0066 |
| t2(MVPA.tmat,MVPA.omat):L.MVPA.42 | -0.0057 | 0.1145 |
| t2(MVPA.tmat,MVPA.omat):L.MVPA.43 | 10.0619 | 0.2981 |
| t2(MVPA.tmat,MVPA.omat):L.MVPA.44 | 0.0442 | 0.1032 |
| t2(MVPA.tmat,MVPA.omat):L.MVPA.45 | 0.2974 | 0.0781 |
| t2(MVPA.tmat,MVPA.omat):L.MVPA.46 | 0.5677 | 0.2809 |
| t2(MVPA.tmat,MVPA.omat):L.MVPA.47 | -0.5547 | 0.0165 |
| t2(MVPA.tmat,MVPA.omat):L.MVPA.48 | 0.6484 | 0.387 |
| s(pfr_month_cos.tmat):L.pfr_month_cos.1 | -0.0001 | 0.2867 |
| s(pfr_month_cos.tmat):L.pfr_month_cos.2 | -0.0001 | 0.2627 |
| s(pfr_month_cos.tmat):L.pfr_month_cos.3 | -0.0002 | 0.623 |
| s(pfr_month_cos.tmat):L.pfr_month_cos.4 | 0.0003 | 0.7492 |
| s(pfr_month_cos.tmat):L.pfr_month_cos.5 | 0.0002 | 0.5422 |
| s(pfr_month_cos.tmat):L.pfr_month_cos.6 | -1.6094 | 1.8354 |
| s(pfr_month_cos.tmat):L.pfr_month_cos.7 | -3.1231 | 5.9218 |
| s(pfr_month_sin.tmat):L.pfr_month_sin.1 | -8.7097 | 71.675 |
| s(pfr_month_sin.tmat):L.pfr_month_sin.2 | -1.0972 | 65.6637 |
| s(pfr_month_sin.tmat):L.pfr_month_sin.3 | 0.0284 | 156.1398 |
| s(pfr_month_sin.tmat):L.pfr_month_sin.4 | 12.0161 | 187.3235 |
| s(pfr_month_sin.tmat):L.pfr_month_sin.5 | 7.3255 | 135.9212 |
| s(pfr_month_sin.tmat):L.pfr_month_sin.6 | 19.7849 | 30.3586 |
| s(pfr_month_sin.tmat):L.pfr_month_sin.7 | 6.1957 | 25.4692 |
| s(pfr_participant).1 | -0.8195 | 3.2219 |
| s(pfr_participant).2 | -3.3994 | 2.33 |
| s(pfr_participant).3 | -0.3429 | 3.0436 |
| s(pfr_participant).4 | -1.9507 | 1.8907 |
| s(pfr_participant).5 | -3.1004 | 1.8541 |
| s(pfr_participant).6 | 2.7715 | 1.7634 |
| s(pfr_participant).7 | -4.7251 | 2.5942 |
| s(pfr_participant).8 | 6.4256 | 2.302 |
| s(pfr_participant).9 | -2.2581 | 1.7001 |
| s(pfr_participant).10 | 2.5425 | 2.4007 |
| s(pfr_participant).11 | -5.2992 | 2.2013 |
| s(pfr_participant).12 | 7.0277 | 3.3273 |
| s(pfr_participant).13 | 2.9095 | 2.0834 |
| s(pfr_participant).14 | 1.1844 | 1.7011 |
| s(pfr_participant).15 | -0.7041 | 3.3855 |
| s(pfr_participant).16 | 3.6399 | 1.6518 |
| s(pfr_participant).17 | 1.327 | 3.2448 |
| s(pfr_participant).18 | 0.2492 | 1.9396 |
| s(pfr_participant).19 | -0.6143 | 1.7548 |
| s(pfr_participant).20 | -3.9002 | 1.8232 |
| s(pfr_participant).21 | -3.2474 | 1.81 |
| s(pfr_participant).22 | 3.3082 | 2.3462 |
| s(pfr_participant).23 | -1.9553 | 2.7352 |
| s(pfr_participant).24 | 0.4892 | 3.3464 |
| s(pfr_participant).25 | -1.0614 | 2.0089 |
| s(pfr_participant).26 | 7.3399 | 1.7229 |
| s(pfr_participant).27 | -2.2067 | 3.3497 |
| s(pfr_participant).28 | -2.5714 | 2.4798 |
| s(pfr_participant).29 | 4.0968 | 2.4943 |
| s(pfr_participant).30 | 3.2534 | 2.0713 |
| s(pfr_participant).31 | -1.0354 | 2.924 |
| s(pfr_participant).32 | 1.6512 | 2.4759 |
| s(pfr_participant).33 | 0.0849 | 1.7427 |
| s(pfr_participant).34 | 0.9219 | 3.3273 |
| s(pfr_participant).35 | -1.5758 | 1.8535 |
| s(pfr_participant).36 | -1.1452 | 2.757 |
| s(pfr_participant).37 | 7.3676 | 2.7764 |
| s(pfr_participant).38 | 0.1432 | 3.0061 |
| s(pfr_participant).39 | -0.5169 | 2.8012 |
| s(pfr_participant).40 | -2.5837 | 3.4357 |
| s(pfr_participant).41 | -2.4264 | 1.8566 |
| s(pfr_participant).42 | 7.4204 | 2.7515 |
| s(pfr_participant).43 | 3.701 | 3.2246 |
| s(pfr_participant).44 | -2.3142 | 2.7271 |
| s(pfr_participant).45 | -1.4783 | 1.8362 |
| s(pfr_participant).46 | -0.8119 | 3.4236 |
| s(pfr_participant).47 | -2.91 | 2.2243 |
| s(pfr_participant).48 | -2.3047 | 2.7677 |
| s(pfr_participant).49 | -1.9364 | 2.7055 |
| s(pfr_participant).50 | -0.8793 | 3.229 |
| s(pfr_participant).51 | -1.9784 | 3.0708 |
| s(pfr_participant).52 | -8.8145 | 1.9939 |
| s(pfr_participant).53 | -0.1424 | 2.7878 |
| s(pfr_participant).54 | -4.7399 | 2.6715 |
| s(pfr_participant).55 | 11.8348 | 1.8121 |
| s(pfr_participant).56 | -0.0268 | 3.1258 |
| s(pfr_participant).57 | -0.9704 | 1.8023 |
| s(week).1 | 0 | 0.0008 |

**Table S3.** Linear predictors for the PFR model.

| **Predictor** | **Estimate** | **SE** | **T.value** | **P.value** |
| --- | --- | --- | --- | --- |
| (Intercept) | 2.9429 | 0.0872 | 33.752 | 0 |
| MPQ-VAS | -1.16 | 0.4964 | -2.3368 | 0.0206 |
| Physical Functioning | 2.2409 | 0.598 | 3.7475 | 0.0002 |
| Psychiatric Diagnosis | 0.2181 | 0.8874 | 0.2458 | 0.8061 |
| Employed | 4.0117 | 1.0939 | 3.6674 | 0.0003 |
| Employed - Other | 6.0273 | 1.2929 | 4.6617 | 0 |
| Divorced | 0.3939 | 1.8963 | 0.2077 | 0.8357 |
| Married | 3.5996 | 0.8582 | 4.1946 | 0 |
| Age | -1.1946 | 0.4629 | -2.5804 | 0.0107 |

